# Gene expression and coexpression alterations marking evolution of bladder cancer

**DOI:** 10.1101/2021.06.15.21258890

**Authors:** Rafael Stroggilos, Maria Frantzi, Jerome Zoidakis, Emmanouil Mavrogeorgis, Marika Mokou, Maria G Roubelakis, Harald Mischak, Antonia Vlahou

**Author notes:** Corresponding author: Antonia Vlahou, PhD, Biomedical Research Foundation, Academy of Athens, Soranou Efessiou 4, 11527, Athens, Greece.

## Abstract

Despite advancements in therapeutics, Bladder Cancer (BLCA) constitutes a major clinical burden, with locally advanced and metastatic cases facing poor survival rates. Aiming at expanding our knowledge of BLCA molecular pathophysiology, we integrated 1,508 publicly available, primary, well-characterized BLCA transcriptomes and investigated alterations in gene expression with stage (T0-Ta-T1-T2-T3-T4). We identified 157 genes and several pathways related prominently with cell cycle, showing a monotonically up- or down-regulated trend with higher disease stage. Genome wide coexpression across stages further revealed intrinsic and microenvironmental gene rewiring programs that shape BLCA evolution. Novel associations between epigenetic factors (CBX7, ZFP2) and BLCA survival were validated in external data. T0 together with advanced stages were heavily infiltrated with immune cells, but of distinct populations. We found AIF1 to be a novel driver of macrophage-based immunosuppression in T4 tumors. Our results suggest a continuum of alterations with increasing malignancy.

## INTRODUCTION

Bladder Cancer (BLCA) accounts for approximately 200,000 annual deaths worldwide and is considered the most expensive cancer to manage[1]. Advances in imaging technologies and drug discovery have improved patient survival and quality of life [1]. However, early-stage incidents [classified as non-muscle invasive (NMI)], still suffer from high rates of disease recurrence, whereas advanced stage [classified as muscle invasive (MI)], and metastatic cancers face poor outcomes [2].

Advances in state-of-art molecular profiling technologies have enabled deeper investigations of BLCA, expanding our current understanding of its molecular pathology. According to the dual track model of bladder carcinogenesis [3], papillary-NMI and MI disease develop from different sets of molecular alterations. Studies performing mutational profiling suggest that low grade Ta tumors arise from activating mutations in either FGFR3 or HRAS, which typically result in over-activation of the downstream Akt/PIK3CA/mTOR and RTK/MAPK growth pathways [3]. In contrast, MIBC tumors are thought to develop from dysplastic Tis lesions with non-functional (mutated) TP53 or RB1 tumor suppressive pathways [3]. However, it remains unclear how these mutational signatures translate to different gene expression routes. Moreover, the dual track model cannot explain adequately molecular events driving transformation of a papillary-NMI tumor to MI, nor in the case of MIBC, alterations happening before the dissemination to detrusor muscle.

In an effort to better understand molecular pathogenesis, various BLCA molecular subtypes have been described [4-15]. Data supporting a continuum of alterations that likely drive bladder carcinogenesis came from MI patients having tumors with a mosaic of both intraepithelial (Tis) and papillary growth patterns, and from MI cancers having traits of papillary-NMI related mutations. Approximately 22% of MI tumors present with activating mutations in PI3KCA and homozygous deletion of the CDKN2A locus, respectively [9], while CDKN2A deletion in MIBC has been observed to occur more frequently in FGFR3 mutated than wild type tumors [16]. Interestingly, comparative mutational analysis between low grade NMI, high grade NMI and MIBC revealed smooth increments or declines in the frequency of mutations in driver genes (FGFR3, KDM6A, TP53, CDKN2A) with increasing malignancy [17]. Additionally, multi-omics analysis of NMIBC identified dysfunctional TP53 and RB1 pathways in about ∼25% of both Ta and T1 tumors [15].

The clinical distinction between NMI and MI diseases and the current understanding of their molecular determinants cannot describe adequately the events driving tumor evolution. On the other hand, molecular subtypes may have important utilities in the diagnosis, prognosis, and decision making, but unfortunately, they represent static entities and their dynamics can only be studied between baseline diagnosis and future recurrences/progressions. In contrast, stage as an ordinal variable reflecting tumor size and depth of invasion, offers a better opportunity for studying the progressive alterations marking tumor initiation, growth, dissemination to detrusor muscle and metastasis.

Gene expression studies comparing the stage profiles of BLCA typically involve small sample sizes and are often limited to comparing NMI and MI. To obtain insight into the trajectories of cancer evolution in BLCA, we collected publicly available transcriptomes from bulk tissue samples, and performed a comprehensive stage analysis of 1,508 subjects with primary BLCA, including a novel integrated pathway-to-network analysis. As the disease progresses gradually to higher stages, our results indicate tumor dependencies on concerted alterations of gene expression, with most prominent those involved in cell cycle regulation.

## METHODS

### Dataset mining

A comprehensive data mining strategy was employed to retrieve studies applying -omics technologies in BLCA. The overall workflow is summarized in **Figure 1**. All genomic urothelial cancer data from cBioportal (including The Cancer Genome Atlas) were downloaded (5/1/2020). Gene Expression Omnibus (GEO) was queried for transcriptomics, additional genomics or protein array datasets using the search terms “bladder cancer” and “urothelial carcinoma”. We also queried ArrayExpress using the special filter “Array express data only” to obtain any additional datasets missing from GEO. All cohort data published or updated between 2010 and 2019, annotated as Homo sapiens, coming from tissue samples with sample size ≥10, were initially retrieved (25/1/2020). All used datasets were published and downloaded anonymized.

**Figure 1:**
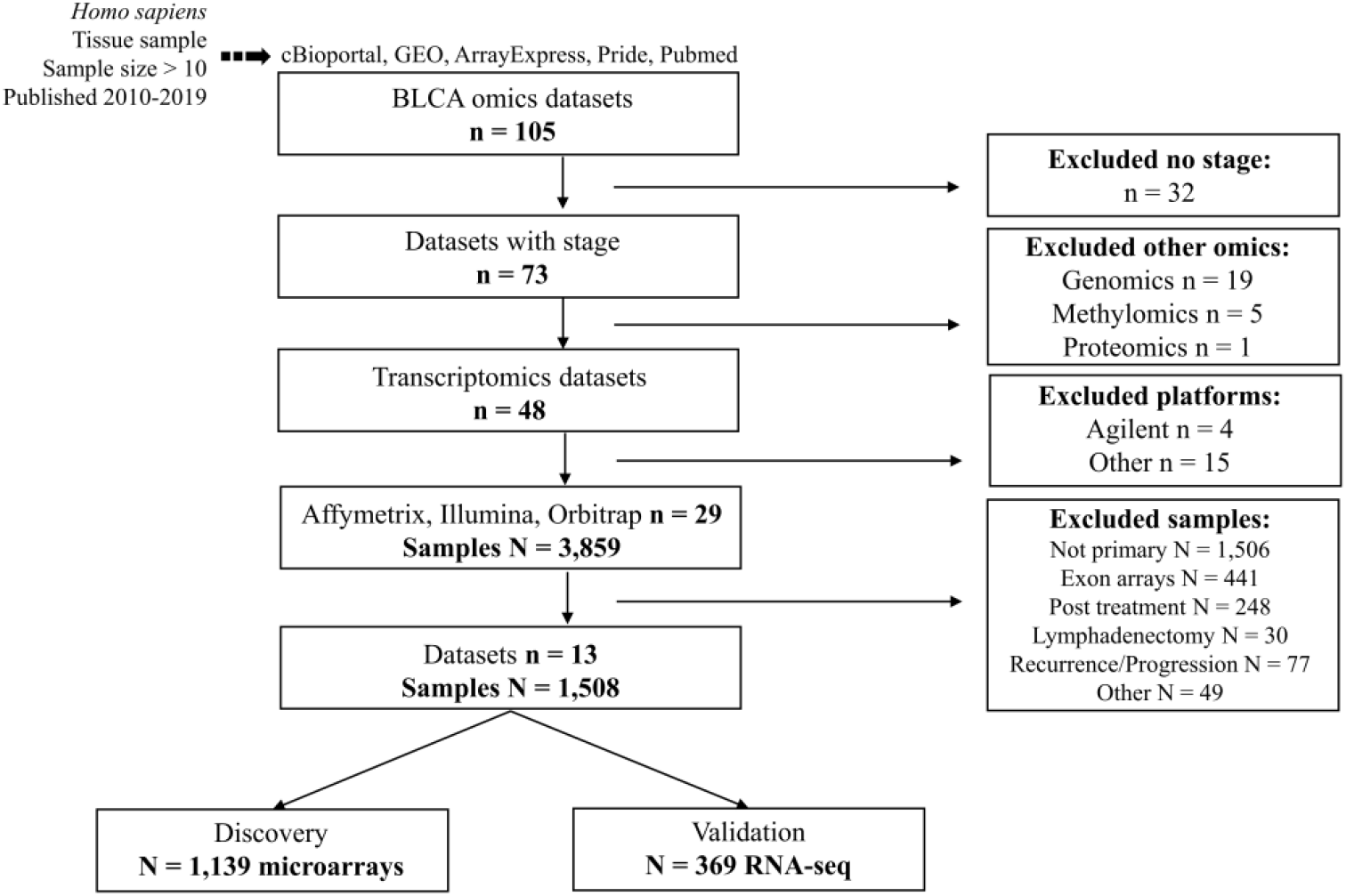
Study design and workflow for the integrative analysis of primary BLCA transcriptomes.

### Integration of the transcriptomes

Microarray data were summarized to gene level with the package *oligo* [18]. Affymetrix were normalized with the RMA method, while Illumina were filtered based on detection p value < 0.05, followed by quantile normalization, addition of 1, and transformation to the natural logarithmic scale. Data were annotated using *biomaRt* (v2.42.1) [19] and mircroarray probes matching to multiple genes were excluded from downstream analysis. The probe with the highest mean across arrays was selected as representative in cases where multiple probes matched to the same gene. Merging of expression matrices was based on the Hugo Gene Symbol using the intersection of genes between studies.

### Adjustment for batch effects

To correct for batch effects across different studies in the discovery data, *ComBat* [20], *removeBatchEffect* [21], and *naiveRandRuv* [22] were evaluated. ComBat performed best and was chosen for further use (manuscript in preparation). The quality of corrected data was assessed with *BatchQC* [23] (**Supplementary Figures 1-2)**, with boxplots of expression distribution per sample and principal component analysis plot of sample relationships, with gene expression comparisons of housekeeping to other genes, and with a set of 12 BLCA markers with known regulation across normal-NMI-MI or across normal-low grade-high grade disease (**Supplementary Figure 3**).

### Differential expression, monotonicity and functional annotation

To identify genes that form a continuum of changes across BLCA stages, each disease stage was compared against normal samples. Genes significantly affected (Mann-Whitney p < 0.05 and had also same orientation of change in all comparisons), were extracted (n = 3,108 genes). We refer to this set as Concordantly Differentially Expressed Genes (CDEGs). Monotonicity for a gene was defined as being a CDEG and additionally having a continuously larger/smaller fold change with increasing stage (fold change, as defined in each disease stage versus normal control). Functional annotation and enrichment were performed using PubMed and the online tool GeneCards (https://ga.genecards.org/), respectively.

### Monotonicity in pathway activation and analysis for gene coexpression

We utilized CDEGs to infer pathway activation scores and to create stage coexpression networks. Pathway activation scores per sample were calculated with the *ssGSEA*-*GSVA* method [24], using the Molecular Signature Database libraries of Hallmark, Canonical Pathways (Reactome subset), C3 (GO biological processes subset) and C5 (GTRD subset of transcription factor targets). *Dorothea* (https://github.com/saezlab/dorothea) [25] was utilized to assess regulon activity. Activation scores across stages were compared with Mann-Whitney tests, and direction of change was defined based on fold change (= Mean of stage – Mean of T0). Pathways showing a monotonal change in the activation scores with increasing stage were extracted. Monotonicity for a pathway was defined similarly to the previous section, i.e. being significantly different in all stage comparisons to T0, and also having a continuously larger/smaller fold change with increasing stage. Stromal infiltration scores were imputed with the ESTIMATE algorithm [26].

Gene-pair coexpression weights (non-negative) were approximated with ensemble learning, using GENIE3 [27] and direction of coexpression (positive/negative) was determined by Spearman’s coefficient. Stage specific networks were constructed with igraph using the top 5,600 positively correlated gene pair interactions with the highest weights for each of the stages. Networks were analysed with Louvain clustering [28] to identify local modules of significantly higher coexpression relatioships (communities) within each stage. The top 5 in size (= number of genes) communities of coxpressed genes per stage were analysed for Gene Ontology Biological Processes [29]. Significance was thresholded at p<0.05 after FDR adjustment according to Benjamini-Hochberg. Hub genes in the monotonal networks were defined based on the betweenness centrality metric.

### Statistical analysis

For continuous variables, significance was defined at Mann-Whitney p < 0.05, unless stated otherwise. Categorical variables were investigated for significance with the Chi-squared test and were adjusted for multiple hypothesis (CRAN package *RVAideMemoire*). All statistical tests were two sided. Gene Ontology and Reactome pathway enrichment analysis was conducted with functions from the package *ClusterProfiler* that performs a two-sided hypergeometric test. All reported correlation scores correspond to the Spearman’s Rank coefficient. Kaplan-Meier analysis and cox proportional hazards regression were performed with the CRAN packages *survminer* and *survival*, and statistical significance was determined with the log-rank method. CIBERSORT analysis was conducted in the web platform https://cibersort.stanford.edu/, and only samples with successful deconvolution (p < 0.05, n = 350 samples) were further used for the comparisons of relative immune populations among stages. All processing, analyses and visualizations were created in the R programming language (version 4.0.2), using predominantly Bioconductor libraries.

## RESULTS

### Cohort description and data quality controls

To perform a comprehensive investigation of publicly available omics studies, we collected 105 datasets comprising more than 8,000 individuals (**Figure 1**). Minimum inclusion criteria for datasets were defined as having at least stage information per subject. To maintain high integrity, we selected microarray data quantified by commercially available single-color channel vendors (Affymetrix and Illumina). Samples or datasets collected after administration of neoadjuvant chemotherapy (NAC) or not clearly annotated as primary were excluded, generating a final dataset corresponding to 1,508 patients. This included in total 12 microarray studies which were employed as a discovery set (**Table 1**) and in addition, the TCGA 2017 BLCA project based on RNA-seq analysis was used as a validation set (**Figure 1**).

**Table 1:** Stage distribution across the microarray datasets used for the discovery phase.

| Technology | Platform | Dataset ID | T0 | Ta | T1 | T2 | T3 | T4 | Tis | Study size |
| --- | --- | --- | --- | --- | --- | --- | --- | --- | --- | --- |
| Affymetrix | GPL17586 | GSE121711 | 10 | 3 | 2 | 3 |  |  |  | 18 |
| Affymetrix | GPL17586 | GSE93527 |  | 79 |  |  |  |  |  | 79 |
| Affymetrix | GPL570 | E-MTAB-1940 | 4 | 41 | 41 |  |  |  |  | 86 |
| Affymetrix | GPL570 | GSE31684 |  | 5 | 10 | 16 | 41 | 18 |  | 90 |
| Affymetrix | GPL6244 | GSE104922 |  | 19 | 7 | 10 | 5 | 0 |  | 41 |
| Affymetrix | GPL6244 | GSE128959 |  | 13 | 39 |  |  |  | 1 | 53 |
| Affymetrix | GPL6244 | GSE83586 |  | 13 | 44 | 241 | 1 | 1 | 1 | 301 |
| Illumina | GPL14951 | GSE48276 |  |  | 2 | 6 | 23 | 6 |  | 37 |
| Illumina | GPL14951 | GSE52219 |  |  |  | 15 | 7 | 1 |  | 23 |
| Illumina | GPL14951 | GSE69795 |  |  | 3 | 7 | 27 |  |  | 37 |
| Illumina | GPL6102 | GSE13507 | 67 | 23 | 80 | 31 | 19 | 12 |  | 232 |
| Illumina | GPL6947 | GSE48075 |  | 33 | 34 | 42 | 23 | 8 | 2 | 142 |
|  |  | **Stage size** | 81 | 229 | 262 | 371 | 146 | 46 | 4 | 1139 |

The compiled discovery cohort comprised 1,058 primary bladder cancer tumor transcriptomes of treatment-naïve patients without prior cancer history, along with 81 adjacent to the cancer bladder samples, a total of 1,139 gene expression profiles (**Table 1**). The ratio of men:woman was 3.5:1, with equal distribution among NMI and MI disease (p = 0.99) and similar mean ages at baseline diagnosis (68 years, p = 0.81). Percentages of normal samples, NMI, and MI in the dataset were *7*.*1%, 43*.*5%*, and *49*.*4%*, respectively (**Table 1**), with the grade distribution being as follows: *16*.*5%* low grade, *48*.*8%* high grade disease with the remaining samples lacking available grade information. Detailed histological records were missing for 71.5% of the cohort, with the most frequently reported histology among the available records being urothelial/papillary (= 23.3%) with squamous differentiation being the most frequent variant (= 1.3%). Annotations included mutation status for FGFR3, PI3KCA, RAS, RB1 and TP53.

Removing batch effects with ComBat was assessed with Relative Log Expression and Principal Component Analysis plots (**Figure 2A**), as well as with comparative analysis of expression levels between houekeeping and non-housekeeping genes (**Figure 2B**), with the algorithm BatchQC (**Supplementary Figures 1 and 2**) [23], or using a set of 12 positive BLCA markers with known regulation across normal-NMI-MI or normal-low-high grade disease (**Supplementary Figure 3**). ComBat successfully eliminated batch effects while maintaining the biological variability (**Figures 2A-C**). BatchQC reports indicated that the variability in the corrected data was generally explained by stage rather than the batch variable **(Supplementary Figure 2)**, allowing for an in-depth downsteam analysis.

**Figure 2:**
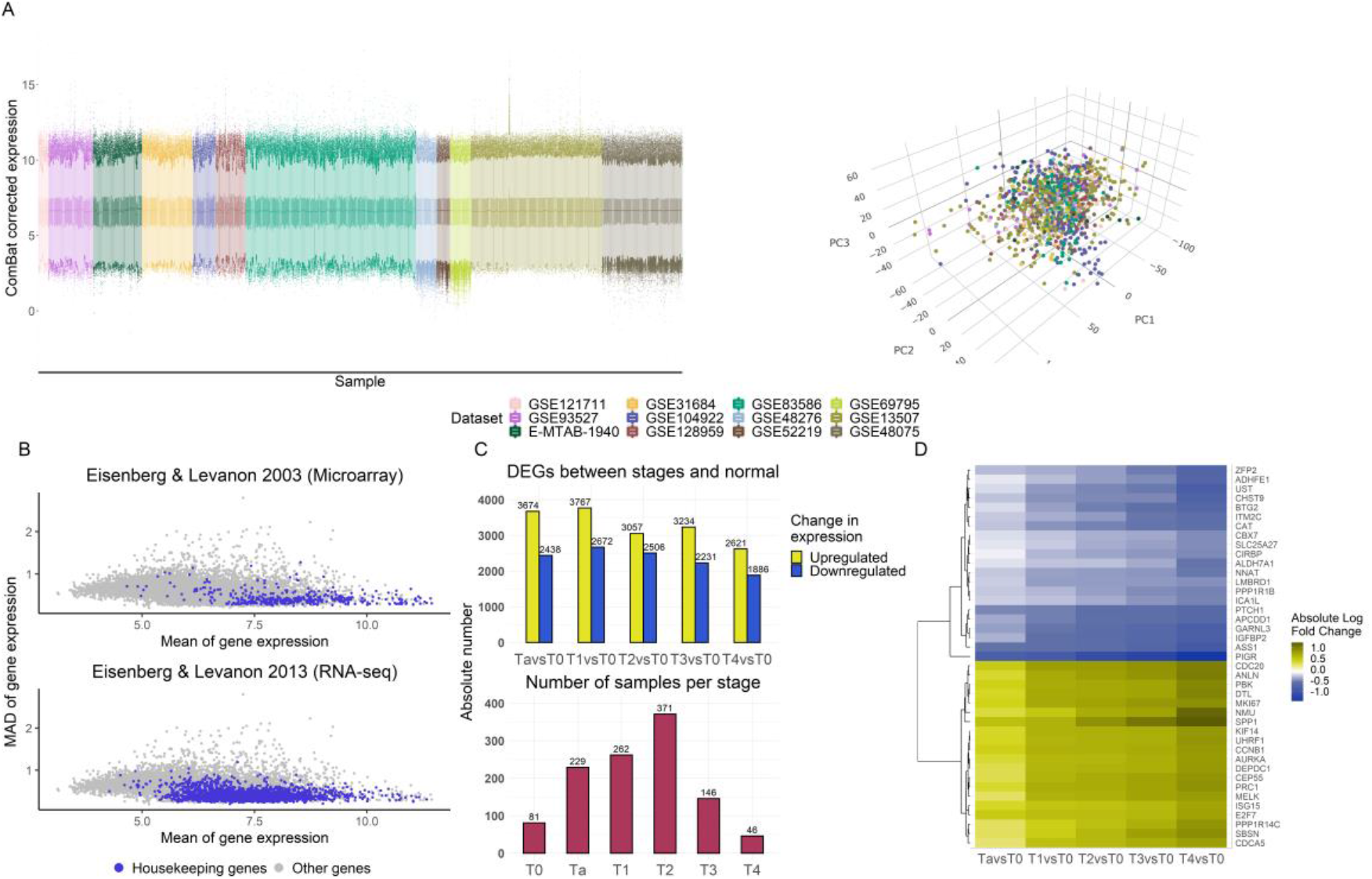
ComBat corrected expression data of the discovery cohort and respective differential gene expression analysis A) Relative Log Expression (right) and Principal Component Analysis plots (left) showing gene expression levels per sample and sample relationships, respectively; the different colors denote the different datasets B) dispersion-to-expression plots illustrating preservation of housekeeping gene properties (higher mean expression and lower median absolute deviation (MAD) compared to non-housekeeping genes) in the corrected data, regardless of microarray (top) or RNAseq (bottom) defined housekeeping genes; C) numbers of differentially expressed genes (DEGs) between disease stages and normal adjacent urothelium (top), along with respective group sizes (bottom), E) heatmap of the top (based on fold change) 20 downregulated and top 20 upregulated monotonically changing genes with increasing disease stage.

### Monotonically changing genes in BLCA regulate growth and cell-cycle progression

For initial assessment of the gene expression relations between normal adjacent urothelium (referred to as T0) and cancerous samples, we performed differential expression analysis of T0 versus NMI and T0 versus MI cancers. When compared to T0, a strong positive correlation (r=0.83, p < 2.1×10^-16) was observed between the two sets of log fold change values, suggesting mutual concordance in abundance change and directionality between NMI and MI tumors. To investigate transcriptional changes associated with increasing malignancy, CDEGs were extracted from the comparisons Ta vs T0, T1 vs T0, T2 vs T0, T3 vs T0 and T4 vs T0. A total of 157 genes were identified having a monotonal (i.e. continuously increasing or decreasing in comparison to T0) change in expression across stages, of which 118 were up- and 39 were downregulated with increasing stage (**Table 2**). Functional annotation revealed that for 46 of these genes, experimental evidence on mediating cell cycle progression exists (Table 2; **Supplementary Table 1**). Upregulated cell-cycle associated genes (n = 44) were not phase specific and included cyclins, DNA polymerases, regulators of the cohesin complex and kinetochore components. The two downregulated cell-cycle associated genes, BTG2 and CDC14B, are both tumor suppressors. The list included 23 genes involved in signal transduction (**Table 2**), 6 of which (ARHGAP11A, AURKA, CDKN3, PBK, PLK1, RRM2), promote cell-cycle progession and were all upregulated with increased stage. The data also indicated an overactivation of the Wnt pathway with increasing disease stage, with its upstream inhibitor APCDD1 being downregulated and its activating ligand WNT2 upregulated (**Supplementarty Table 1**). Fourteen of the 157 genes were transcriptional or translational regulators (**Table 2**), including genes with known upregulation in bladder cancer (transcription factors E2F1, DEPDC1 [30, 31]). Based on the monotonal changes with increasing stage, increased androgen receptor activity may be predicted, as both its translational enhancer BUD31 [32] and its downstream transcritption factor ELK1 [33] were upregulated. Four (HTR2C, LRP8, NENF, NMU) of the 157 genes are involved in neurotransmission or neuronal development, all upregulated (**Table 2**). Among the 157 genes, 21 genes were of not well described or unknown function (**Supplementary Table 1**), including the oncogenic factor TRIM65 [34] found upregulated with bladder cancer stage. Additionally, upregulation was detected for the cisplatin resistance gene CLPTM1L, as well as for SRD5A1, which catalyzes the conversion of testosterone and progesterone to their 5-alpha forms [35]. Further functional enrichment using GeneCards for the 157 genes verified their involvement in cell-cycle pathways, with top hits being related to the regulation of the Anaphase-promoting (APC) complex (score = 31.53), to PLK1 (score = 24.47) and Aurora B (score = 20. 95) signaling, as well as to TP53 (score = 19.06) and RB1 (score = 17.85) cell cycle checkpoint control (**Supplementary Table 2**). Using the same tool, analysis for compounds with potentially therapeutic benefit provided as top hit the DNA alkylating agent Bendamustine (score = 19.23) (**Supplementary Table 3**) [36]. Univariate cox regression analysis indicated 29 genes with potentially prognostic impact at p < 0.01 (**Supplementary Table 4**).

**Table 2:**
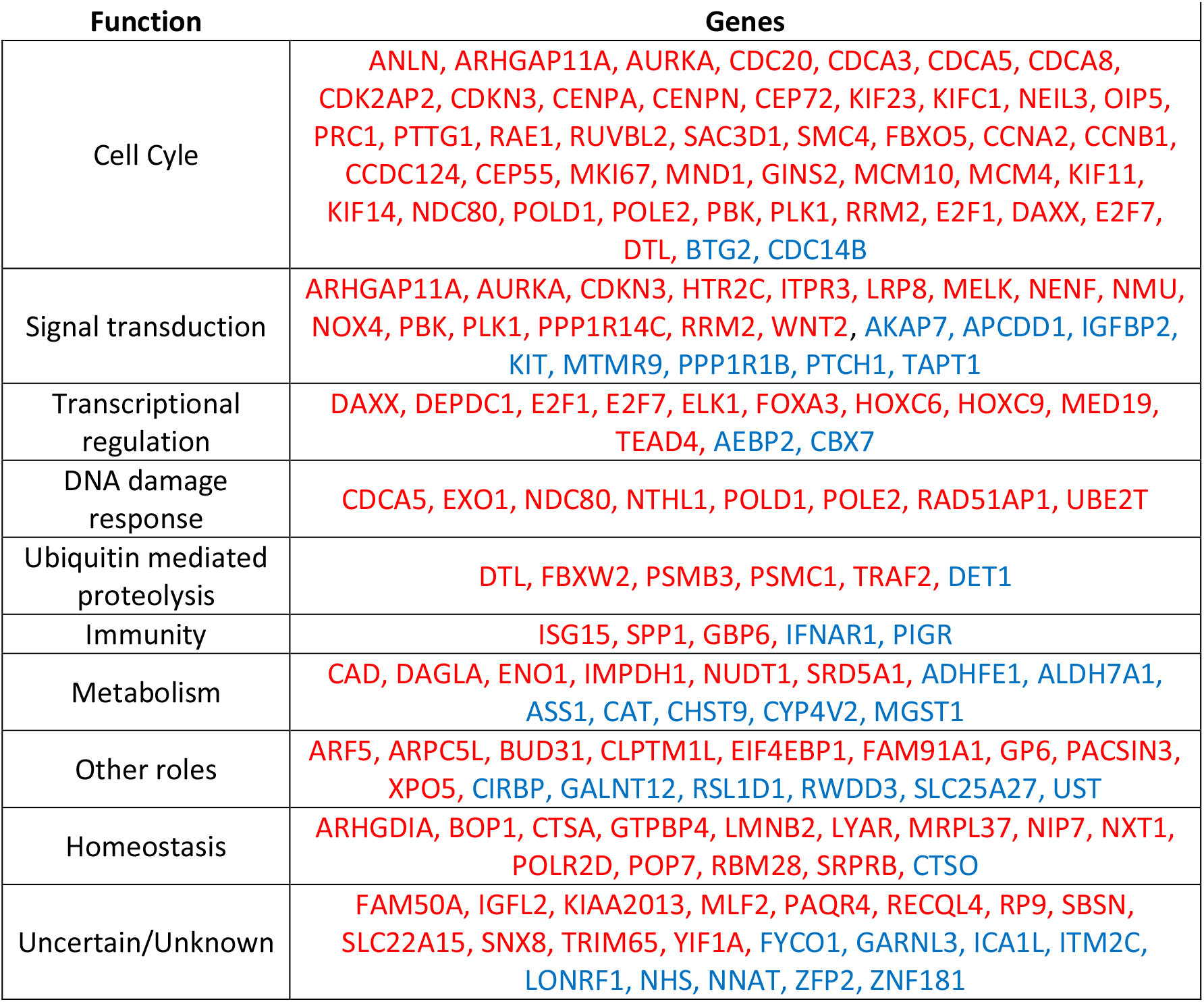
Classification of the 157 monotonically changing genes with increasing disease BLCA stage to functional categories. Red and blue colors correspond to upregulation and downregulation, respectively. More details per gene are provided in **Supplementary Table 1**.

### The expanded dataset of 3,108 CDEGs links cell-cycle, ECM remodelling and metabolic alterations with increasing disease stage

To increase coverage of the BLCA alterations, we further extracted Concordantly Differentially Expressed Genes (defined in Methods; CDEGs, n = 3,108) from the comparisons of cancerous stages to normal adjacent urothelium and investigated the differentially activated pathways they represent, using ssGSEA. Similar to the analysis of monotonicity, we sought to identify pathways with monotonically up- or down-regulated activation scores across T0 and BLCA stages. Results indicated gradually stronger activations of several mitotic processes, positive regulation of the canonical Wnt pathway, mTORC1 signaling, expression of MYC targets, degradation of anaphase inhibitors, metabolism of nucleotides, mobility of formins, and the TNFR2/non-canonical NF-kB pathway (**Figure 3, Supplementary Table 5**). Conversely, consistently reduced expression versus controls was observed in genes involved in lipid and fatty acid catabolic processes, in the metabolism of heme, in pathways promoting muscle cell differentiation and, interestingly, in genes regulating the circadian clock (**Figure 3**). Regulon activity per sample was estimated and respective scores between cancerous stages and normal tissue were compared. GATA3 and GLI2 were the only regulons that significantly associated with increasing malignancy with their activity being downregulated (**Figure 3**).

**Figure 3:**
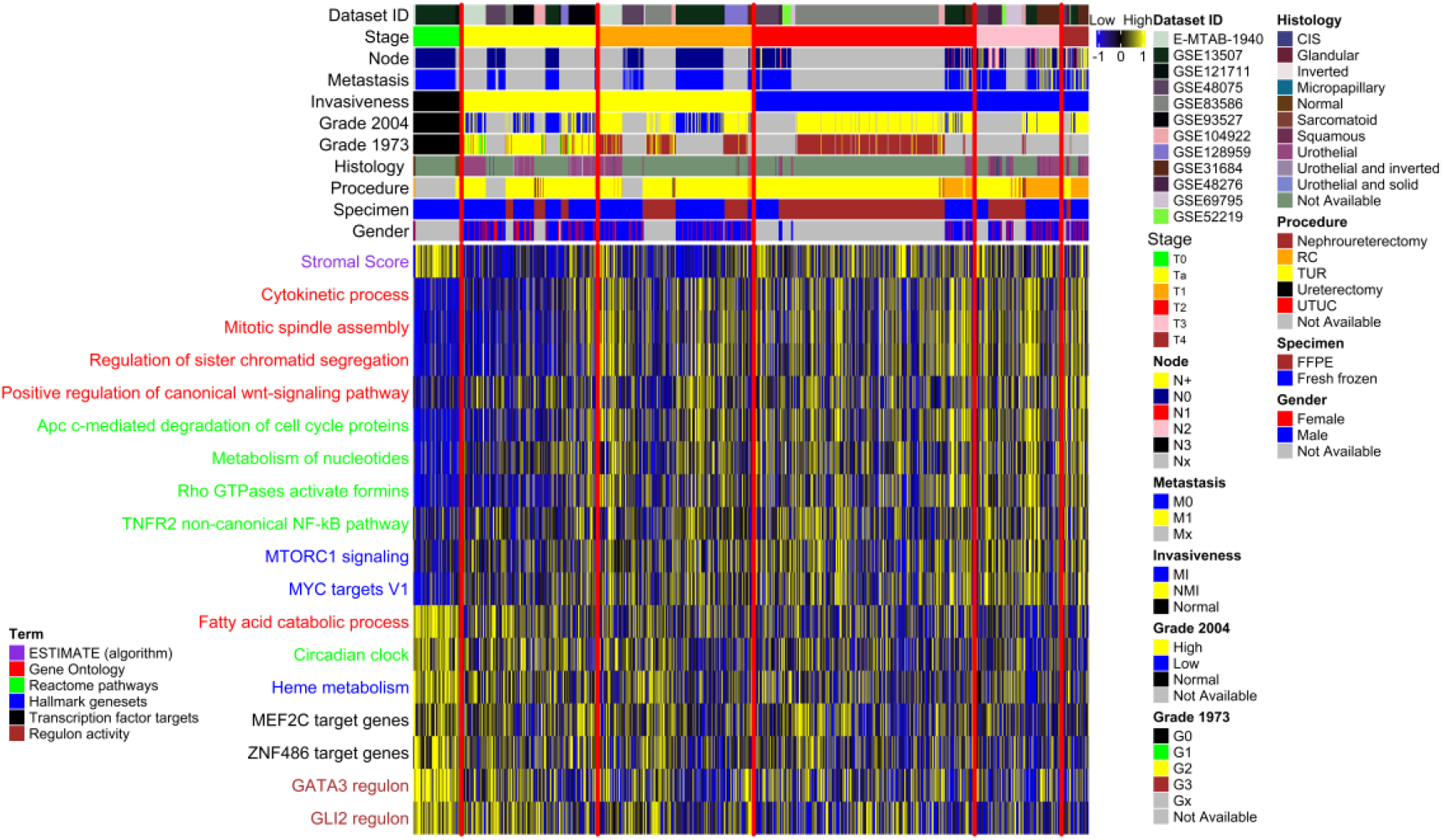
Excerpt of the pathways showing a monotonal increase or decline in their activation scores with higher stage.

To further investigate genome wide alterations and coexpression patterns associated with cancer progression, an integrated network-pathway analysis of the different disease stages was performed, using the 3,108 CDEGs. Out of 9,659,664 weighted gene pair interactions, the top 5,600 were used as the most representative to build the corresponding networks, applying Louvain clustering to identify topologically connected communities (modules). Retrieved gene clusters have high degree of coexpression, and thus changes in their composition across stages reflect gains/losses in pathway rewiring. We used the Gene Ontology – Biological Process library to identify molecular processes affected by changes in coexpression inside the top 5 largest in size (based on number of genes) communities (**Figure 4, Supplementary Tables 6-11**). The analysis revealed large differences in gene coexpression between T0 and cancerous samples with 4 out of the 5 largest in size communities associating clearly with specific biological processes.

**Figure 4:**
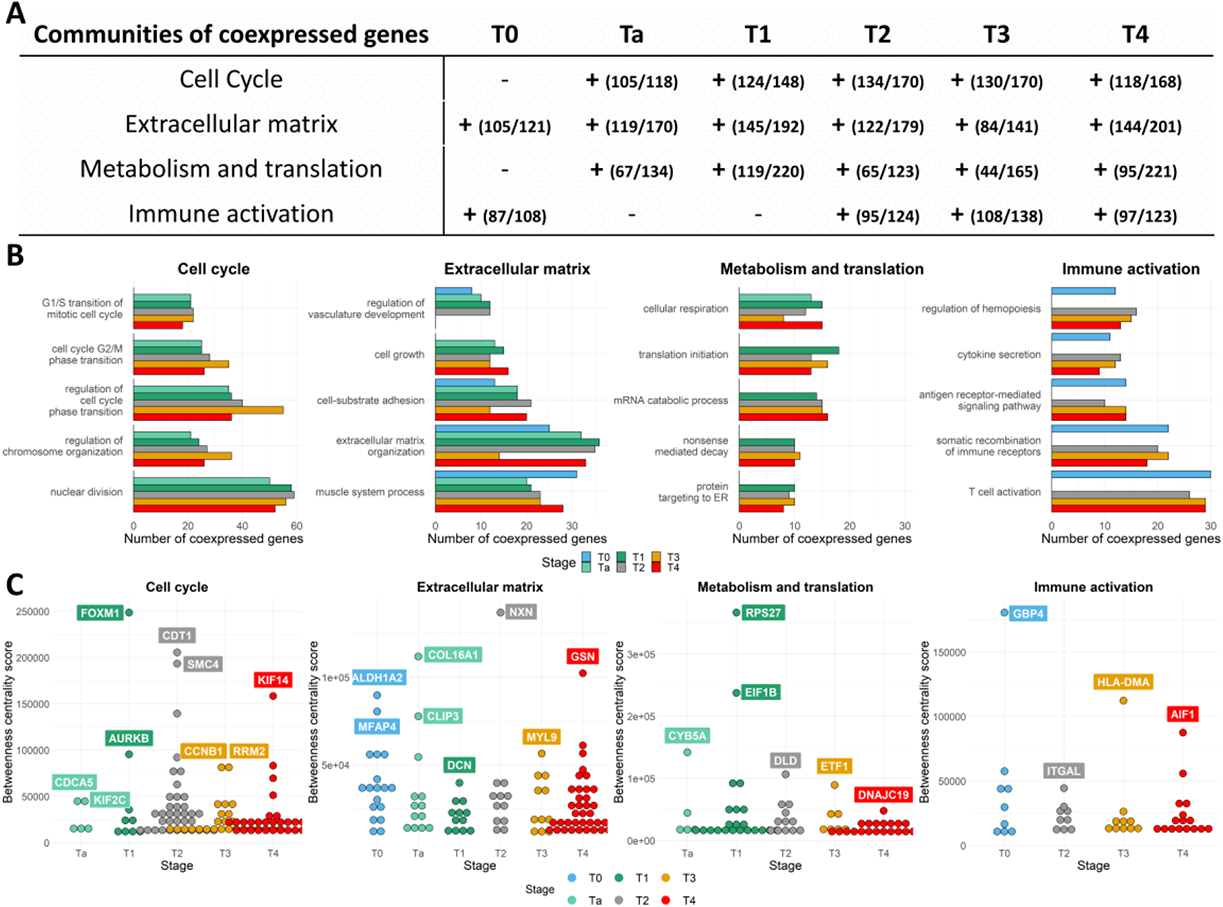
Biological process analysis of the largest in size coexpressed communities identified in each BLCA stage, using the 3,108 CDEGs between T0 and cancerous stages. **A)** Coherent communities identified and characterized across T0 and disease stages. Numbers in parentheses show the fraction of genes with existing Biological Process annotation with respect to the total number of genes found to be coexpressed in each of the community **B)** Barplots of the most significantly enriched biological processes per community depicting number of coexpressed genes for each. **C**) Hub genes identified across the studied conditions based on the betweenness centrality scores (y axis).

Three out of the top 5 communities were consistently detected in all BLCA stage networks. Based on examination of their enriched processes, these were labeled as 1) the cell-cycle community, 2) the ECM and developmental community 3) the metabolic and translational community (**Figures 4A, 4B**)

When investigating the cell cycle community and specifically the processes of G1/S transition of mitotic cell cycle, cell-cycle G2/M phase transition, regulation of cell cycle phase transition, regulation of chromosome organization and nuclear division, most of the participating genes were found to be coexpressed in all BLCA stages (**Figure 4B**). However, the gene size of the cell cycle community increased with higher stage (Ta n = 118, T1 n = 148 and MIBC n = 168-170), and interestingly, the proportion of the genes with unknown or unclear function (lacking GO annotation) also increased (11.9% for Ta, 29.8% for T4). Among the genes with cell cycle annotation (n=184), 80 genes were coexpressed in all cancerous stages and were also upregulated in malignancy compared to T0, possibly forming the backbone of cell cycle progression in BLCA (Supplementary Table 13). However, only 36 of them had a monotonal increase in the expression levels with higher stage (Supplementary Table 13). To detect potential drivers of variation in the cell cycle co-expression profiles of stages, we looked into the betweenness centrality scores of the genes inside each of the stage cell cycle communities (Figure 4C), and found CDCA5 and KIF2C to be hub genes in Ta tumors, FOXM1 and AURKB in T1, CDT1 and SMC4 in T2, CCNB1 and RRM2 in T3 and KIF14 in T4 tumors (Figure 4C).

The community of ECM and developmental processes was enriched in cell-cell communication and cell-matrix interactions, in responses to microenvironmental stress, as well as in differentiation programs of epithelial, mesenchymal and stem cells (**Figure 5B**). The biological process of extracellular matrix organization included coexpressions of 15-36 genes of which COL13A1, FGFR4, FOXF2 and SCUBE1 were coexpressed only in T0 compared to disease stages, whereas 26 genes (including COL6A1/A2, COL16A1, MFAP5, MMP11) were coexpressed in malignancy but not in T0. In line with recent observations [37], we noticed that T0 presented with an active ECM remodeling profile. Sixteen of the ECM associated genes were coexpressed both in T0 and in the NMIBC stages, including genes promoting ECM degradation and tumor cell migration (MMP2, CTSK, PDPN), fibrotic collagens (COL1A2, COL6A3, COL14A1, COL15A1), and pro-angiogenic factors (PDGFRA, RECK). Notably, coexpression in the T0 samples was predicted to be driven by the cancer stem cell marker ALDH1A2 and MFAP4 (**Figure 4C**), while in Ta tumors, by COL16A1 and CLIP3. Since CLIP3 interacts with both AKT1 and AKT2 [38], CLIP3 may have an important role in the early AKT/PI3K/mTOR axis of carcinogenesis. For the same process of extracellular matrix organization, compared to Ta, T1 tumors had gains in ADAMTSL2, while compared to T1, T2 tumors had gains in MMP11 and LRP1. Similarly compared to T2, T3 tumors had coexpression gains in ITGA10 and in genes of the endocytic pathway (ABL1, CYP1B1) whereas compared to T3, T4 tumors had gains in DDR2 and JAM2.

The community of metabolism and translation encompassed mitochondrial, translational and multiple metabolic processes being activated during carcinogenesis, and was more profound in the T1 and more advanced tumors. The processes of cellular respiration, translational initiation, mRNA catabolic process, nonsense mediated decay and protein targeting to ER were consistently enriched in most of BLCA stages. The results highlighted a set of 12 genes commonly co-expressed across stages for these processes, including COX7B, DLD, NDUFS4, UQCRFS1, PAIP2, RPL15, RPL30, RPL7, RPS23, RPS27, RPS27A, RPS4X, with RPL36A and YTHDF3 being T2-specific, COQ10B, EIF4E3, MTIF2, NCPB1 as T3-specific while EIF4E1, ISCU and GSPT2 were T4-specific.

Besides the abovementioned consistently detected communities in BLCA, a community enriched in processes of immune cell activation and cytokine secretion, was identified in the T0 and the MIBC stages and involved both innate and adaptive responses, as well as, processes of immune cell adhesion and migration. It was the least variable community with respect to different processes and respective genes coexpressed across the different stages. Strikingly, T0 and the MIBC stages had similar profiles. Out of the 17 genes of the process of T-cell activation that were commonly coexpressed at the MIBC stages, 15 were also coexpressed in the T0 samples. To further investigate these observations, the transcriptome data per sample were deconvoluted into relative abundances of immune cell populations using CIBERSORT, and cell fractions between disease stages were compared. Significant results were obtained for the following populations: CD8+, activated CD4+, activated NK, Monocytes, Macrophages M2 and activated Dendritic cells (**Supplementary Figure 4**). Results indicated differential commitment of immune cells to T0 and BLCA stages. T0 (n = 37) were significantly more infiltrated with CD8+ (p = 0.046) and monocytes (p = 4.6E-4) than tumor samples (n = 313), verifying the presence of the immune community in the T0 samples. However, compared to tumor, T0 samples had significantly less abundance of activated CD4+ cells (p = 5.28E-3), of macrophages (p = 16.8E-5), of activated dendritic cells (p = 0.0002) and of activated natural killer cells (p = 0.015). Generally, NMIBC had lower infiltration burden than MIBC, while compared to other BLCA stages, Ta tumors (n = 34) were significantly more infiltrated with activated dendritic cells (p = 0.024). Abundance of CD8+, of activated NK cells, and of M2 macrophages increased linearly with higher malignancy. Interestingly, AIF1 a gene that promotes macrophage survival [39], was found to be driver of immune coexpression in the T4 tumors.

### RNA-seq independent analysis validates presence of communities and prognostic genes

In lack of another dataset comprising all the disease stage spectrum of primary BLCA incidents, the observed alterations in the discovery set were investigated for their reproducibility in the TCGA-BLCA publication [9]. Although the validation set is not perfectly suited since it includes only stages T2-T3-T4, we used it due to the RNA-seq technology which offers a good reliability at the validated findings. Differential expression analysis among the available stage comparisons (T3 vs T2 and T4 vs T3) in the TCGA data validated 74 of the 157 monotonal genes in the comparison T3vsT2, and none in the comparison T4vsT3 (**Supplementary Table 13**). Cox regression analysis of the 157 genes in the TCGA data validated 12 genes having a prognostic value (**Supplementary Table 4**), including IMPDH1, MRPL37, MED19, ENO1, ANLN, GTPBP4, MLF2, higher levels of which associating with worse survival and CBX7, ZFP2, AKAP7, ICA1L, CDC14B higher levels of which associating with better survival probability. To validate as possible the coexpression analysis findings, stage specific coexpression networks were also created using the TCGA data, and were clustered with the Louvain algorithm. GO-Biological process analysis of the communities validated the differential segregation of the cell-cycle, extracellular matrix and immune activation processes to distinct communities (**Supplementary Figure 5**).

## DISCUSSION

Early integration of raw BLCA data has been previously performed, in the context of characterizing molecular subtypes [40], or validating results of either scRNA-seq [41] or RNA-seq re-analysis [42]. In this study we performed an early integration meta-analysis of normal looking urothelium and BLCA stages, aiming to identify continuous gene expression alterations with increasing malignancy. To our knowledge, this is the first attempt to associate molecular alterations with clinical classification, by analyzing more than one thousand well-characterized, primary samples. Instead of focusing onto molecular subtypes, we increased power and addressed the disease as a continuum, under the assumption that individual samples reflect different snapshots of the whole process. Our novel design based on the hypothesis of continuous evolution through stages, has been successfully applied here and resulted in novel findings on gene regulation associated with cancer progression.

First, we identified a set of 157 genes with a monotonal change in expression across T0 and BLCA stages. Almost half of these genes were components of the cell cycle machinery, or kinases signaling positively for it, or transcription factors responsible for the expression of cell cycle genes. Twelve of the 157 genes had also prognostic value in external RNA-seq data. Of these, apart from ENO1 higher levels of which had been previously linked with worse BLCA outcome [43], all the remaining 11 associations to survival are novel. IMPDH1 plays a role in cancer progression by promoting cell growth, and higher expression levels of this gene have been observed in MIBC compared to normal tissue [44]. Similarly, MED19 a component of the mediator complex that regulates the transcription of RNA-polymerases, was found overexpressed by IHC in human BLCA compared to normal tissues, and its knockdown in the T24 and 5637 bladder cell lines resulted in cell-cylce arrest at the G0/G1 checkpoint and also in attenuation of cell growth [45]. The involvement of MRPL37, GTPBP4 and MLF2 in BLCA development has not been characterized, but oncogenic properties have been attributed to these genes in other contexts. ANLN and AKAP7 are thought to regulate bladder cell growth and apoptosis in a TP53 independent manner [46]. ICA1L is naturally expressed in sperm cells and its implication in BLCA has not been investigated. The CDC14B gene is located on the 9q chromosome, a region that is often deleted in BLCA. This might also explain its downregulation in malignancy, as observed in the discovery set. CDC14B is believed to dephosphorylate TP53 [47], but the functional consequence on the mitotic or DNA damage repair pathways is not yet well clarified [48]. CBX7 is a component of the chromatin modifier PRC1-complex and is required for the propagation of the transcriptionally repressive state of multiple genes through cell-division during embryonic development [49], including Hox genes [50]. Expectedly, we noticed that HOXC6 and HOXC9 were both monotonically upregulated with increasing malignancy (**Supplementary Table 1**). ZFP2 is a probable transcription factor and evidence suggest an epigenetic role as well [51], while recently, high load of mutations in another member of the ZFP family (ZFP36) were associated with upper tract urothelial carcinoma [52].

Our analysis highlighted expected landmark pathways, such as mTORC1 pathway [53] and MYC targets [54] which were upregulated, but also novel downregulated pathways such us the circadian clock and the metabolism of heme. These results associate for the first time BLCA progression to the disruption of the circadian homeostasis and to iron metabolism deficiencies, events that are thought to be tumorigenic [55, 56], but their exact mechanism of action is not well understood. We detected an “incomplete” Warburg effect in which the monotonical downregulation (or deactivation) of lipid and fatty acids metabolism, was not accompanied by any upregulation of glycolysis. Additionally, to the GATA3 regulon which is a known driver of luminal biology, we detected a novel progressive downregulation of the GLI2 regulon. GLI proteins are transcription factors of the Sonic hedgehog (Shh) pathway and although GLI2 expression levels positively correlate with more invasive BLCA cell lines, Shh genes do not behave accordingly [57]. Our results validate these observations, as the entire regulon of the GLI2 TF was inactivated with increasing BLCA malignancy, suggesting no potential therapeutic effect in its inhibition.

A change in the expression levels of a single gene may not always have a functional consequence, mainly due to the complementary action of other genes of that pathway. Instead, a concerted change in the expression levels of multiple genes is often required for the cell to successfully transit between phenotypes. We investigated this type of changes using a network coexpression analysis followed by intra-stage graph clustering, and were able to detect both intrinsic and microenvironmental gene rewiring programs that shape tumor evolution. These gene programs were found to be organized into distinct biological communities (which were validated in the TCGA 2017 study), and consisted of genes stably coexpressed among all BLCA stages, but also of genes specifically coexpressed (gained or lost) in a particular stage only. We assume that the ECM remodelling relies mostly on this last property of gains and losses in coexpression, since the bladder consists of heterogeneous anatomical tissues with stiff biological barriers (basement membrane, muscle layer), which in turn require specialized cell functions to be dissolved. The immune activation community was present in both T0 and MIBC. Results of the CIBERSORT analysis showed that inside the bladder tumor and with increasing stage, most monocytes preferentially differentiate into macrophages with M2 polarization. This is in line with the recent findings from Chen et al. [41] in which authors analyzed scRNA-seq data of BLCA patients and observed a similar pattern of differentiation for monocytes. As the CIBERSORT algorithm has not been designed to recognize signatures of T-cell exhaustion, CD8+ cells appeared at higher number in T0 samples compared to tumor, however we cannot exclude the existence of biological barriers in the normal adjacent urothelium that forbid CD8+ cells from reaching tumor. T4 samples had the highest percent of M2-macrophages, which could partially explain their immunosuppressive state, and as a novel finding, T4 coexpression in the immune cells was driven by AIF1, a gene that ensures the survival and proliferation of macrophages. We hypothesize that immune evasion in BLCA is likely promoted by M2-macrophages that actively express AIF1, but further work is required to validate these observations.

Our study has its limitations, which include the retrospective nature of the analysis, and the usage of batch correction methods to integrate different microarrays. The batch correction method unavoidably eliminates some of the biological variability leading to increased false negative rate; our validation of the applied method via checking expression of known genes, in combination to focusing on positive signals, suggests that sufficient biological variability is maintained. In addition, restrictions in the validation set imposed by lack of samples from all disease stages did not allow validating the observations particularly at the NMIBC. Finally, clinical stage is known to have varying rates of misdiagnosis. However, the high number of samples used in each of the stage categories is expected to balance out to some extent misdiagnosed cases while increasing power of the received results.

## Data Availability

Data availability
The datasets generated during and/or analysed during the current study are available from the corresponding author on reasonable request.

## Code availability

All custom R scripts created for analysis and visualizations are available by the first and the corresponding author upon reasonable request.

## Data availability

The datasets generated during and/or analysed during the current study are available from the corresponding author on reasonable request.

## Acknowledgements

MM is supported by the European Union’s Horizon 2020 research and innovation programme under the Marie Sklodowska-Curie grant agreement No 898260 (H2020-MSCA-IF-2019, ReDrugBC, Grant agreement ID: 898260).

## Contributions

HM and AV conceived and designed the investigation, RS, MF, MZ and EM processed and analyzed data, RS and EM wrote software for analysis and visualizations, RS, MF, MZ HM and AV interpreted results, RS wrote the manuscript, MM, MR, HM and AV provided critical revisions.

## Ethics declarations/ Competing interests

HM is the co-founder and co-owner of Mosaiques Diagnostics, MF, EM, and MM are employed by Mosaiques Diagnostics.

