## Supplementary Figures for "Gene expression and coexpression alterations marking evolution of bladder cancer"

**Supplementary Figure 1:** Batch effect in the non-corrected data (before adjusting with ComBat)

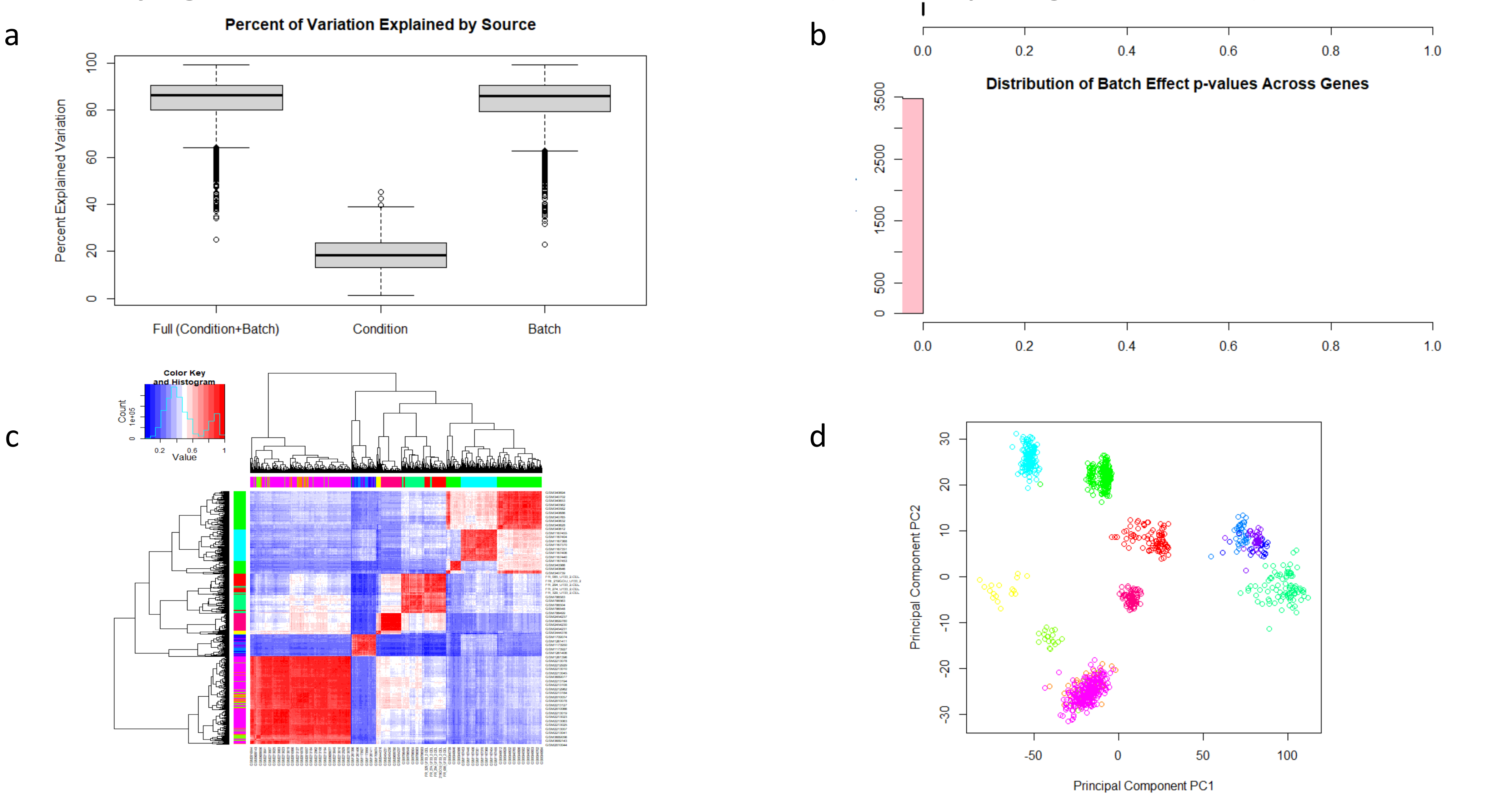


a) Variation explained by clinical stage (Condition) and datasetID (Batch), b) fraction of genes affected by batch (all genes have $p < 0.05$ meaning that they are significantly affected by batch), c) Heatmap of pearson correlation coefficients between sample pairs (column and row colors correspond to the 12 datasets), d) First two principal components showing sample relationships (colors correspond to the 12 datasets)

**Supplementary Figure 2:** Batch effect in the corrected data (after adjusting with ComBat)

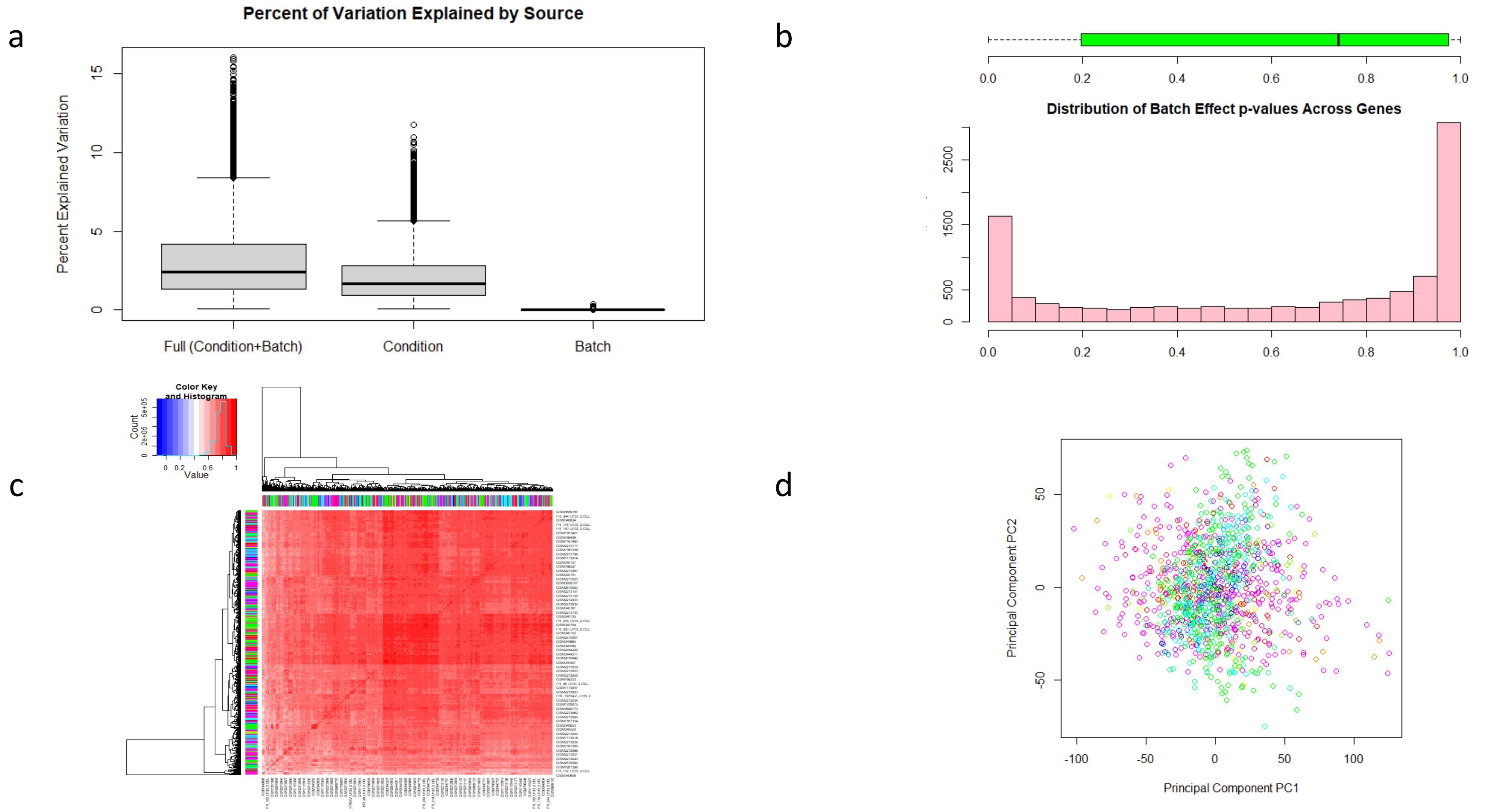


a) Variation explained by clinical stage (Condition) and datasetID (Batch), b) fraction of genes affected by batch (most genes have $p > 0.05$ meaning that they are not significantly affected by batch), c) Heatmap of pearson correlation coefficients between sample pairs (column-row colors correspond to the 12 datasets), d) First two principal components showing sample relationships (colors correspond to the 12 datasets)

**Supplementary Figure 3:** Gene expression of 12 known BLCA markers among clinical conditions in the ComBat corrected data

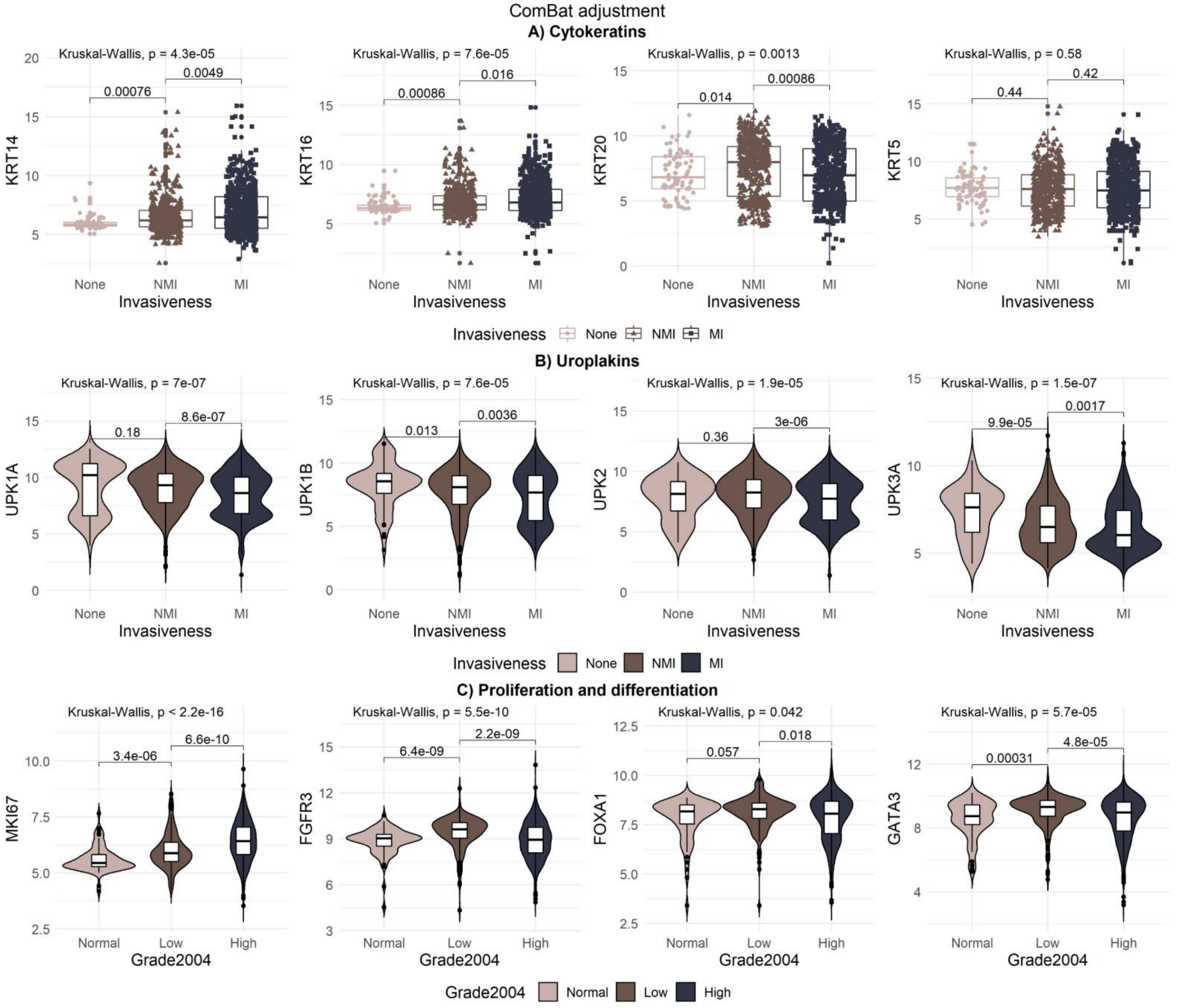

**Supplementary Figure 4:** CIBERSORT analysis of the expression data

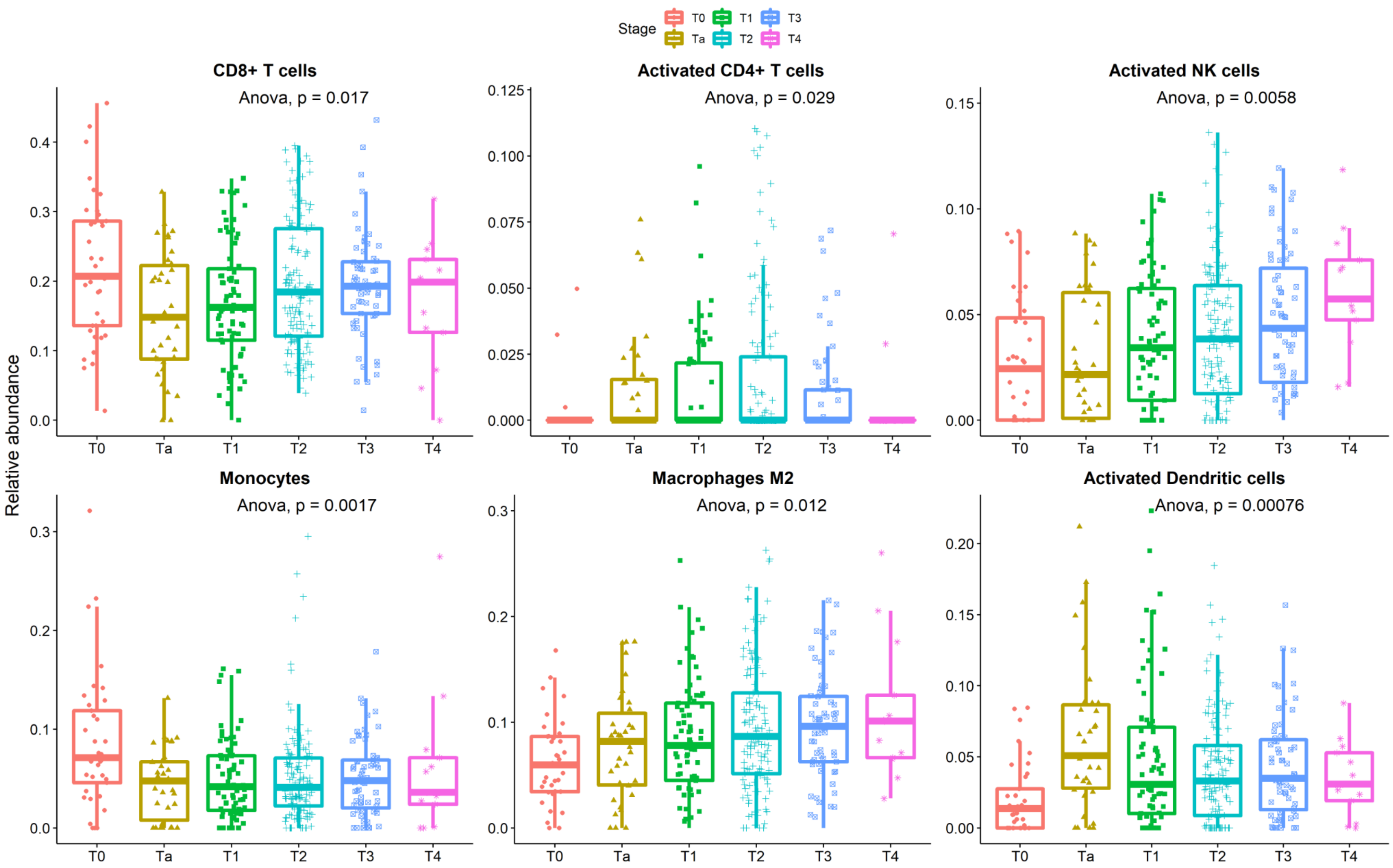

**Supplementary Figure 5:** Coexpression network analysis in the TCGA2017 data and the segregation of coexpression to communities

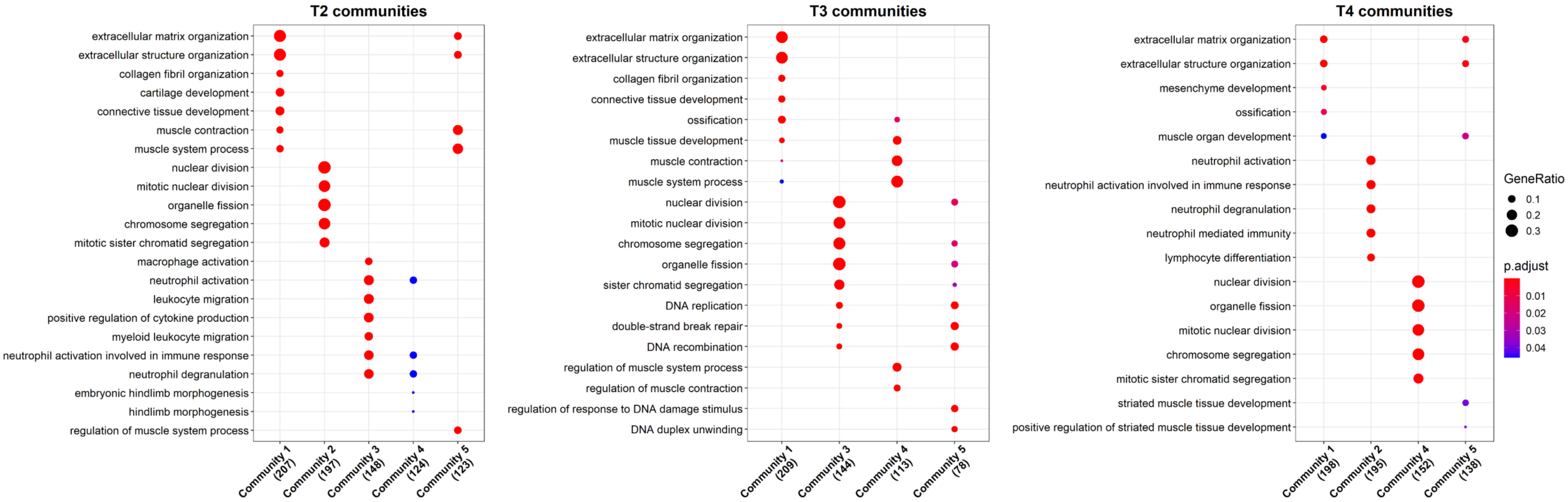
